# Barriers and Facilitators to Facility-Based Feeding Support for Small and Sick Newborns in Ethiopia: A Qualitative Study of Clinicians’ Perspectives

**DOI:** 10.1101/2024.12.19.24319282

**Authors:** Yihenew Alemu Tesfaye, Moses Collins Ekwueme, Heran Biza, Zerihun Tariku, Mulusew Lijalem Belew, Meseret Asefa, Destaw Asnakew, Abebe Gobezayehu, Melissa Young, John Cranmer

## Abstract

Breastfeeding is recognized as the optimal form of infant nutrition. The World Health Organization recommends initiation of breastfeeding within the first hour after birth and exclusive breastfeeding (EBF) for the first six months. However, facility-based breastfeeding practices, especially for small and sick newborns (SSN), face numerous challenges. The Saving Little Lives (SLL) program in Ethiopia seeks to improve SSN survival by promoting comprehensive neonatal healthcare practices, including appropriate feeding. Despite these efforts, limited data exist on clinicians’ experiences and perspectives regarding SSN feeding support in healthcare facilities. This qualitative study explored the neonatal feeding experiences of clinicians in selected SLL facilities in the Amhara region, Ethiopia. Semi-structured interviews revealed multilevel barriers influencing facility-based SSN feeding practices. These barriers were categorized as: (1) facility factors—including insufficient clinician training, staff shortages, and inadequate feeding tools; (2) neonatal/maternal factors—such as health complications in newborns and mothers and maternal concerns about insufficient milk production; and (3) sociocultural factors— including traditional practices like uvulectomy and prelacteal feeding. A key facilitator identified was the role of predominantly female clinicians with personal breastfeeding experience, which positively influenced feeding support efforts. The findings suggest that many barriers are modifiable through targeted interventions, including enhanced clinician training, integration of infant feeding counseling into prenatal and postnatal care, and improved access to feeding tools in healthcare facilities. These insights offer critical guidance for developing evidence-based strategies to strengthen facility-based SSN feeding support, contributing to improved neonatal health outcomes in low-resource settings.

**KEY MESSAGES:** *What is already known on this topic:* Breastfeeding is crucial for infant nutrition, with early initiation and exclusive breastfeeding (EBF) for six months recommended by the World Health Organization. However, challenges persist in supporting small and sick newborns (SSN), particularly in low-resource healthcare settings.

*What this study adds:* This study highlights multilevel barriers to facility-based SSN feeding in Ethiopia. The barriers include clinician training gaps, inadequate feeding tools, insufficient facility-based neonatal feeding support to struggling mothers and neonates, and sociocultural practices like uvulectomy and prelacteal feeding. A key facilitator was having predominantly female clinicians with personal breastfeeding experience.

*How this study might affect research, practice and policy:* The findings inform targeted interventions, such as clinician training, integration of newborn feeding counseling into maternal care, and improved access to feeding tools, shaping research, policy, and practice to enhance neonatal feeding outcomes in low-resource settings.

## INTRODUCTION

In the past decade, significant progress has been made in reducing under-five mortality, but the decline in neonatal mortality has been much slower [1]. Globally, neonatal deaths account for 45% of all under-five deaths, with many of these fatalities linked to complications associated with low birthweight (LBW) [2, 3], particularly affecting SSN [4, 5, 6]. To reduce neonatal mortality and enhance the long-term health of infants, targeted healthcare programs and services for SSN are essential. Extensive evidence shows that facility-based early postnatal interventions—such as Kangaroo Mother Care (KMC), immediate and EBF, and early detection and treatment of suspected infections—can significantly improve the health and survival of SSN [7, 8, 9, 10].

In Ethiopia, the neonatal mortality rate is 33 deaths per 1,000 live births [11]. About 10% of newborns are preterm, and 20% are classified as LBW [12, 13]. Both LBW and SSN are significant contributors to neonatal mortality in the country [14]. Recognizing LBW and SSN as critical public health issues, the Ethiopian government has developed and implemented a national strategy for newborn and child survival, along with targeted interventions to improve outcomes for SSN [15, 16, 17].

The SLL program is one of the Ethiopian government’s interventions being implemented in four targeted regions, in collaboration with local and international partners. Its goal is to reduce SSN mortality by enhancing health system factors and services that address perinatal emergencies and LBW mortality. Supporting SSN through kangaroo mother care (KMC) services is a key component of the SLL minimum care package [18].

While SSN often face challenges with early breastfeeding, experience separation from their mothers, or receive insufficient milk after emergencies, there is currently limited information on targeted interventions to promote EBF or breastmilk feeding. This study aims to provide formative data on global solutions to these feeding challenges by exploring the experiences of clinicians in SLL facilities in the Amhara region of Ethiopia. We focus on their knowledge, practices, support systems, and challenges. The findings will contribute critical evidence for targeted nutrition interventions for SSN, both globally and in Ethiopia.

## METHODS

### Study design, study settings and participants

We conducted a qualitative study to assess facility readiness to provide nutritional support for SSN. Participants were selected from facilities implementing Ethiopia’s SLL program in Amhara, reflecting the clinical diversity of government hospitals, including both primary and referral facilities.

Ethical clearance and a support letter were obtained from the Amhara Public Health Institute (APHI). Clinicians were recruited after the research coordinator presented the APHI support letter and IRB clearance to each facility’s medical director, who informed potential respondents about the study’s objectives. With hospital administration’s permission, clinicians involved in neonatal health services and infant feeding in KMC and/or NICU wards, who consented to participate, were invited to join the study.

### Data collection

The study was conducted from June to July 2022. Our interview guide included questions about clinicians’ roles, facility SSN feeding practices, challenges, proposed solutions, and available resources. Developed by the research team, the guide was pre-tested on a sample of clinicians outside the main study and modified based on team feedback.

Interviews were conducted in person by two trained social science experts using a semi-structured format. Interviews lasted 45 minutes to 1.5 hours and was conducted in Amharic. All interviews were audio recorded.

### Data analysis

The audio-recorded interviews and field notes were translated into English and entered into MAXQDA 2022 Software. We employed thematic and content analysis, using both predetermined themes from existing literature and inductive themes that emerged during coding. YAT developed and revised the codebook with input from the research team. Initial coding was performed by YAT, while MCE double-coded 20% of the interviews to assess inter-coder agreement. We explored patterns and relationships between codes to develop broader themes, which were reviewed by the team for accuracy.

## RESULTS

We interviewed eleven clinicians from seven facilities in the West Gojjam and South Gondar zones of the Amhara region, Ethiopia. The clinicians included trained nurses providing health services to SSN in NICU and/or KMC wards, with experience ranging from 2 to 9 years. Eighty percent of the clinicians were female, and one served as the coordinator for the NICU and KMC wards (Table 1).

**Table 1:** Characteristic of participating clinicians.

| Participant characteristics (n=11) | No. and frequency (%) |  |
| --- | --- | --- |
| **Gender** |  |  |
| Female | 9 (82) | 140 |
| Male | 2 (18) |  |
| **Role** |  |  |
| NICU/KMC coordinator | 1 (9) | 141 |
| Clinicians | 10 (91) |  |
| **Type of facility** |  |  |
| Primary hospital | 5 (71) | 142 |
| General/referral hospital | 2 (29) |  |
| **Years of experience working in NICU/KMC** |  |  |
| 0 to 4 years | 6 (55) | 143 |
| 5 to 10 years | 5 (45) |  |
| **Education** |  |  |
| BS in general nursing | 4 (36) | 144 |
| BS in neonatal nursing | 7 (64) |  |
|  |  | 145 |

We first describe current SSN feeding practices, then outline barriers and challenges to facility-based neonatal nutrition and highlight clinician-implemented solutions to improve feeding support in the study facilities.

### Facilities’ neonatal feeding practices

Newborns admitted to the NICU include both preterm and term infants with health complications, as well as LBW (<2000 gm at birth) infants with feeding difficulties. Very low birthweight (<1500 gm at birth) preterm newborns are typically referred to the NICU for “maintenance IV fluids” until they are ready to feed. In contrast, KMC wards accommodate LBW preterm or term newborns without severe health issues and who can feed, often transferred from the NICU after stabilization.

In KMC wards, clinicians assist with feeding by helping mothers with direct breastfeeding or expressing breastmilk for cup or nasogastric tube (NGT) feeding. If a preterm or LBW neonate cannot feed, they may be readmitted to the NICU for “maintenance IV fluids” until they can feed. A NICU and KMC coordinator summarized their approach:

> We have three activities related to newborn feeding: First, if a newborn cannot feed, we admit them to the NICU for maintenance IV fluids. Second, if a newborn can feed but is low birthweight with no severe health issues, we place them in the KMC room with their mother. Lastly, if the mother struggles to produce enough milk, we support her to breastfeed, and if that remains insufficient, we may resort to formula feeding as a last option.

Four-fifths of the interviewed clinicians were unfamiliar with their facilities’ neonatal feeding care plan or related protocols. Those who did and didn’t acknowledged the existence of such plans described various feeding practices for LBW preterm and term neonates, as well as for very low birthweight (VLBW) neonates.

All clinicians stated that they advise mothers to breastfeed or feed their neonates breastmilk immediately after birth. However, when asked about breastfeeding protocols, clinicians primarily discussed their facility-based practices. They indicated that their initial workflow involves evaluating the newborns’ condition to determine whether to admit them to KMC or NICU or provide necessary support before sending them home. Figure 1 illustrates the facilities’ workflow immediately after birth.

**Figure 1.**
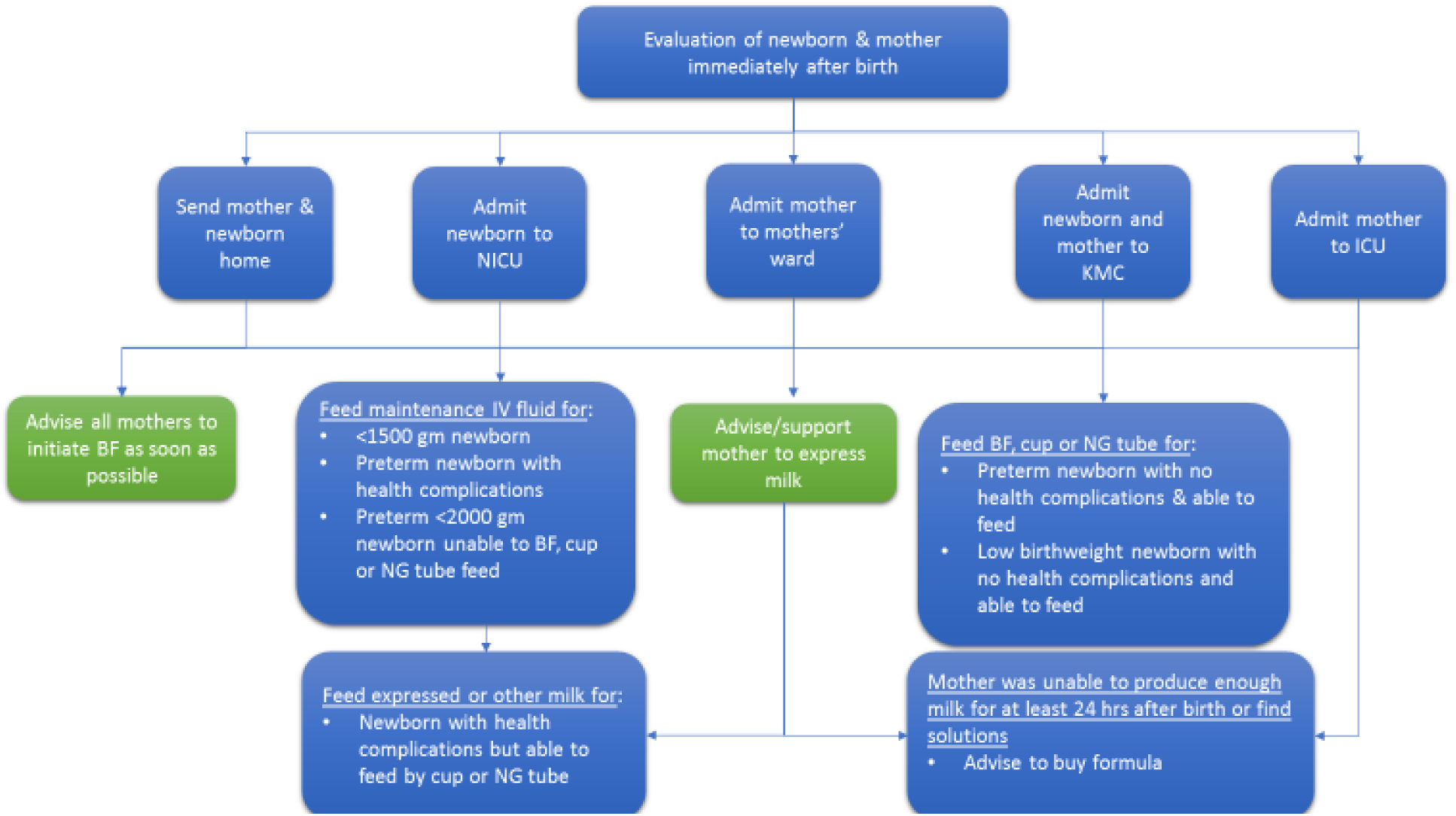
Healthcare facilities’ post-childbirth workflow, highlighting newborn feeding support services.

Clinicians indicated that both the condition of the newborn and the mother jointly influence facility-based feeding practices (Table 2).

**Table 2:**
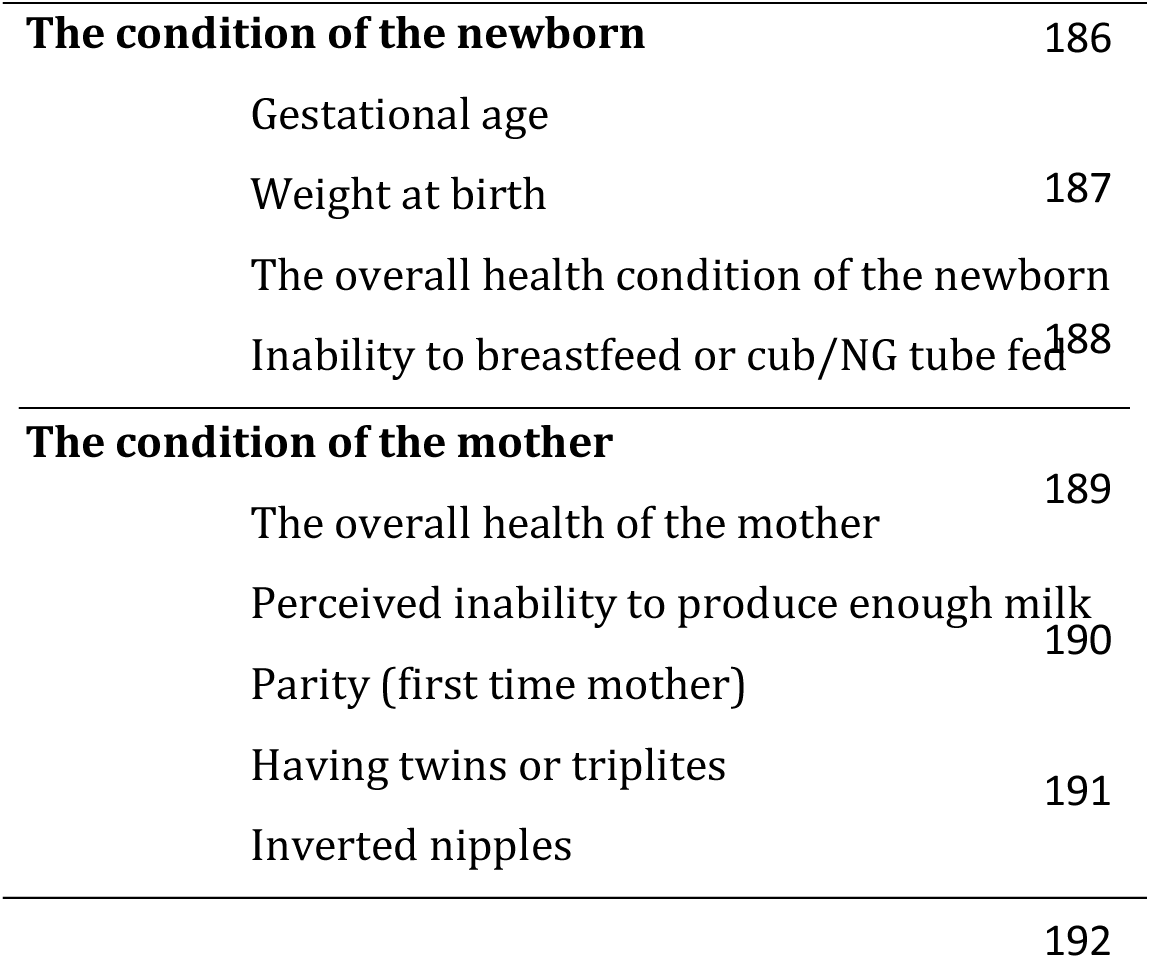
Factors determining facility-based newborn feeding practices.

When discussing factors that influence facility-based feeding practices, one clinician from a referral hospital stated:

> There are several factors that affect our choice of newborn feeding practices. For instance, if a mother is in critical condition, we cannot breastfeed the newborn. Additionally, if the mother has inverted nipples, this also impacts feeding practices.
>
> The health condition of the baby is another important factor; sometimes sick newborns are unable to suckle, so we may use alternatives such as maintenance IV fluids to provide nutrition.

Facilities can have different feeding practices for LBW term and preterm neonates. In addition to differences in concentrations of fluids in the “maintenance IV fluid”, preterm LBW neonates get more feeding attention and support than term LBW neonates because of their development.

There are also different feeding plans for LBW and VLBW neonates. Most VLBW newborns, born too early to feed, are placed in the NICU in an incubator for two to three weeks. In contrast, LBW neonates have a greater chance of staying in KMC, where they can receive breastmilk either through direct BF or expressed milk via cup or NGT.

As part of the SLL project implemented in the study facilities, all participants confirmed that their facilities provide KMC (and NICU) services for SSN. These services aim to improve survival through a package of targeted interventions, including nutritional support. However, when prompted for details about feeding support provided during KMC and NICU services, most clinicians focused more on practices unrelated to newborn feeding.

### Barriers and challenges to facility-based newborn feeding and some solutions

The factors influencing facilities’ feeding support for SSN were multidimensional and encompassed a variety of challenges which were categorized into three main themes: health system (facility) factors; neonatal/maternal factors; and sociocultural factors (Table 3).

**Table 3:** Factors influencing facility-based feeding support for SSN.

| Main themes | Subthemes | Primary codes |
| --- | --- | --- |
| Facility | Staffing & training | Lack of trainings specific to neonatal feeding and clinicians' workload |
|  | Unfamiliarity with BF policy and protocols | Clinicians unfamiliar with facilities' BF policy and protocols |
|  | Resource limitations | Limited resources: space (NICU/KMC rooms) and tools (breast pumps and infant formula) |
| Neonatal/maternal | Neonates' inability to breast feed | Neonates' not able to breastfeed or fed by cup/NGT |
|  | Mothers' condition | Mothers' health condition after birth<br>Mothers' perceived inability to produce enough milk |
|  | Mothers' previous experiences | Mothers' parity (e.g., first time mother)<br>Previous feeding experiences<br>Inverted nipples |
| Sociocultural | Uvulectomy | Cutting neonates' uvula at home |
|  | Prelacteal feedings | Butter feeding to newborns at home |

#### Facility factors

The three subthemes categorized as facility factors include: clinicians’ lack of training specific to neonatal feeding and staffing shortages; clinicians’ unfamiliarity with facility breastfeeding policies and protocols; and limited resources, such as insufficient space and equipment needed to support feeding, including breast pumps.

Only one clinician received training specific to newborn feeding, which was through the SLL project. Another clinician mentioned having NICU care training that included some basics on infant feeding. The remaining informants had no specific training on neonatal feeding, aside from limited education during nursing school. Additionally, clinicians identified staffing shortages as a challenge, as it limits the time available to support neonates and their mothers.

When asked about their facility’s breastfeeding policy and its communication by senior staff, 82% of the clinicians reported being unaware of such policies. One of the two clinicians who were aware noted that the policy focused on “raising awareness about the importance of newborn breastfeeding and the value of having healthy babies for the country’s future.”

Shortages of resources were identified as significant barriers to effective newborn care, including insufficient supplies for neonatal feeding and overcrowded NICU and KMC rooms. Clinicians reported that evaluating newborns and making admission decisions was challenging, especially as many facilities were operating at full capacity including space and experts. Due to these limitations, clinicians often had to prioritize the most at-risk neonates for NICU or KMC admission.

In addition to limited space and personnel, clinicians reported insufficient resources, such as breast pumps, to support newborn feeding. They also noted the unavailability of formula in their facilities and its unaffordability for most families as barriers to using breastmilk substitutes (BMS) when necessary.

When clinicians consider providing BMS for SSN as a last resort, families are typically asked to purchase and provide the formula. However, most families cannot afford it, creating challenges for both the families and the clinicians. One nurse highlighted the unaffordability of BMS for most families, saying:

> In my area, many mothers cannot afford formula when breastfeeding is difficult. If a mother can’t provide enough milk, or the newborn can’t suckle or has health issues, we admit the newborn to the NICU, supplementing maintenance IV fluids for 24 hours while encouraging the mother to express and supply breastmilk. However, if the mother still can’t supply breastmilk, we can’t feed the newborn. Most families can’t afford formula, leading to many newborns dying from hypoglycemia.

Clinicians discussed solutions to some facility-related challenges. One initiative, in collaboration with social workers, aimed to provide essential resources, like formula, for mothers who couldn’t afford them. In a few facilities, when BMS like formula are necessary, clinicians can purchase formula for newborns receiving additional neonatal care, while mothers are encouraged to supply breastmilk.

#### Neonatal/maternal factors

Neonatal and maternal factors were identified through three subthemes related to the conditions of neonates and mothers: the inability of neonates to breastfeed or be fed by cup/NGT; mothers’ health post-birth and perceived inability to produce enough milk; and parity/previous maternal experience. Clinicians highlighted that mothers’ perceived inability to produce breastmilk and neonates’ inability to suckle were significant barriers to neonatal feeding. One clinician noted,

> The second issue, after mothers’ inability to produce enough milk, is that the newborn may not be able to suckle, which is a major obstacle that requires NICU admission.

Clinicians noted that when newborns are unable to suckle, the primary solution is to admit them to the NICU for at least 24 hours of “maintenance IV fluids,” while advising mothers to provide breastmilk for feeding via cup/NGT. If a mother is critically ill and cannot supply breastmilk, clinicians will use BMS and require family members to purchase formula.

#### Sociocultural factors

The final barrier theme identified was unsafe socio-cultural practices. In Amhara, as in many regions of Ethiopia, it is customary for newborns to undergo uvulectomy from traditional health practitioners. Some parents may provide their newborns with prelacteal fluids such as butter at home. Clinicians noted that these harmful practices can complicate neonatal feeding, as many infants admitted after such procedures develop complications. One clinician remarked:

> Despite our recommendations against uvulectomy and butter feeding, some mothers still give butter or allow uvulectomy, complicating our feeding support when they are later admitted.

These practices often occur after discharge or for homebirth newborns, influencing how clinicians manage care and feeding for SSN infants.

## DISCUSSION

The study aimed to identify barriers and challenges to facility-based feeding support for SSN in the Amhara region of Ethiopia, addressing a global gap in evidence-based feeding practices. It revealed several factors influencing neonatal feeding support and highlighted measures clinicians have taken to overcome these challenges.

As part of neonatal care practices, clinicians evaluate the health of the newborn and mother immediately after birth to determine whether to admit or discharge them. Standard procedures dictate that all VLBW newborns and low LBW newborns with health complications including feeding difficulties are admitted to the NICU, while LBW and preterm newborns without health issues are placed in KMC. Neonates without health concerns are discharged. Additionally, VLBW, LBW, and newborns with complications are referred from lower-level facilities, such as health centers, to higher-level facilities at birth.

Barriers to facility-based nutritional support were categorized in to three themes: facility factors, neonatal/maternal factors, and sociocultural factors.

### Facility factors

Facility factors limiting feeding support for SSN included clinicians’ lack of training in newborn feeding, staff shortages, unfamiliarity with the facility’s neonatal feeding policy, and limited resources such as breast pumps and newborn formula. Respondents identified inadequate nutritional training, staffing issues, and resource gaps as the primary barriers to SSN feeding.

Lack of targeted training in neonatal feeding, particularly for SSN nutrition, was identified as a primary barrier to effective feeding support for SSN. Observations and discussions with clinicians and NICU/KMC coordinators revealed that most clinicians received only a one-time comprehensive training on neonatal health. While these trainings covered some basics, clinicians felt they lacked in-depth education on neonatal nutrition. Additionally, the study found that clinicians were unfamiliar with any newborn feeding policies or protocols at their facilities, indicating a communication gap regarding nutritional training and feeding practices from supervisors to front-line clinicians. This gap is further compounded by the lack of targeted nutritional training for SSN care.

Following gaps in training, limited staffing was the second facility factor impacting feeding support. Staffing shortages required clinicians to provide care for multiple neonates and mothers, reducing the time available for supporting SSN. This finding aligns with studies from other settings, where nursing shortages were identified as a significant barrier to effective neonatal feeding practices [19, 20].

Clinicians’ ability to provide lactation support to mothers with SSN is affected by the availability of BF support materials and tools. For instance, when direct breastfeeding isn’t possible or neonates are in the NICU, expressed breastmilk can be offered. However, the lack of breast pumps and insufficient training in hand expression make this challenging.

This can lead clinicians and mothers to consider purchasing supplemental formula, as lactation management issues significantly influence the decision to supplement [21]. Improving access to breast pumps and providing training on hand expression would help address these challenges.

In our context, the unavailability and unaffordability of formula in many facilities posed a significant challenge for rural families in Amhara. Formula supplementation was sometimes deemed essential due to maternal-newborn separation and insufficient support for establishing breastfeeding. For instance, in cases where a financially limited individual gave birth to twins or triplets but couldn’t produce enough milk, formula supplementation became necessary. However, this is complicated by facilities that do not stock formula. While donor human milk could be a solution, it is only viable where safe and affordable milk-banking conditions exist [22].

### Neonatal/maternal factors

Neonates’ inability to feed and various maternal factors—including health conditions, perceived inability to produce sufficient milk, and rare issues like inverted nipples—were key influences on facility-based nutritional interventions for SSN.

Neonates’ inability to suckle during breastfeeding or cup feeding can be influenced by various factors, including health complications or being VLBW [23]. Such neonates are admitted to the NICU and given “maintenance IV fluids” according to facility protocols.

Additionally, mothers’ lack of BF experience with SSN neonates and clinicians’ limited feeding support further restrict newborn suckling.

Maternal factors that decreased SSN feeding support included health conditions after birth, being a first-time mother, and rare cases like inverted nipples. These were the most significant maternal influences on facility-based feeding. Similar to this study, other research has also identified first-time motherhood as a factor affecting successful breastfeeding [21]. Clinicians’ limited knowledge of targeted feeding interventions for SSN, as well as strategies to support first-time mothers or address issues like inverted nipples, can further restrict facility-based feeding support.

In our study, mothers’ perceived inability to produce enough milk was identified as a key factor limiting facility-based breastfeeding. This finding aligns with several studies from various contexts, which also highlighted inadequate milk production as a common concern [24, 25, 26, 27], especially among mothers of preterm and LBW neonates [26, 27].

### Sociocultural factors

Certain sociocultural practices can be harmful and further limit breastfeeding. Parents and traditional healthcare providers often perform uvulectomies and prelacteal feeding for newborns, typically at home after discharge from facilities [28, 29, 30]. Many of these infants later require readmission to hospitals due to complications from these interventions.

Prelacteal feeding, defined as any fluid other than breastmilk given to newborns before breastfeeding begins, is practiced in many regions worldwide [28, 31]. In Ethiopia, 25.5% of newborns receive prelacteal feeds, often consisting of boiled water or butter, with a third being butter [28]. Newborns exposed to these practices may develop infections or gastrointestinal complications, leading to hospital referrals. Such situations are particularly challenging due to barriers in facilities, including low staffing, limited protocols, and inadequate clinician skills for targeted nutritional support.

In Amhara, uvulectomy is a common traditional remedy for uvula inflammation, for example in Gondar city 84.3% of newborns under six months undergo the procedure [29]. This practice often leads to complications requiring hospital admissions, hindering clinicians’ ability to provide effective neonatal care and nutrition support.

### Implications

This study identified three key factors influencing facility-based feeding support for SSN. While categorized separately, understanding their interconnectedness is essential for improving feeding outcomes. For instance, clinicians’ unfamiliarity with neonatal feeding policies can stem from a lack of specific training, affecting their ability to support mothers and newborns facing breastfeeding difficulties. Additionally, staffing shortages compel clinicians to assist multiple mothers simultaneously, reducing the time available for adequate education and support, which in turn impacts mothers’ breastfeeding challenges. Similarly, a lack of breastfeeding support tools limits clinicians’ ability to provide effective lactation support and affects mothers’ perceptions of their milk supply.

To promote long-term change, clinicians suggested several strategies to enhance facility-based feeding support for SSN. Regular training specific to SSN feeding support, along with consistent communication of neonatal feeding policies, can boost clinicians’ capacity and confidence. This can be achieved through tailored on-the-job and standard training covering proper breastfeeding practices, breastmilk expression techniques, and nutrition.

Developing and distributing educational materials and counseling tools on neonatal nutrition and proper breastfeeding practices can further improve health outcomes. Increasing access to essential tools, such as breast pumps, will also facilitate feeding support. While budgeting and staffing shortages are challenging, identifying breastfeeding champions among existing staff and promoting ongoing initiatives can help mitigate resource constraints and enhance SSN feeding outcomes.

## LIMITATIONS AND STRINGTHS OF THE STUDY

To our knowledge, this is the first study to investigate clinicians’ experiences with SSN feeding in Ethiopian facilities. By exploring experiences across various tiers of Ethiopia’s health services continuum, we documented a range of insights from the Amhara region. While our sample was drawn from a subset of government hospitals in Amhara and may not represent all facilities and clinicians in the region, in-depth interviews with diverse clinicians helped mitigate potential biases, such as social desirability. Although our smaller sample size could also be seen as a limitation, data saturation was achieved, and the rich qualitative data collected through semi-structured interviews and participant observations provided a comprehensive understanding of the issues. Additionally, our research assistants, with extensive qualitative experience and no direct ties to Emory-Ethiopia or the study facilities, ensured an impartial data collection process.

## CONCLUSION

Our findings highlight that the factors contributing to facility-based SSN feeding are multifactorial but can be addressed through targeted interventions. Key steps to enhance SSN feeding support include improving clinicians’ capacity through on-the-job and standard training programs; developing and distributing tailored neonatal nutrition educational materials and counseling tools; and providing perinatal counseling sessions to mothers on neonatal nutrition and proper breastfeeding practices. Additionally, enhancing facilities’ access to the necessary tools for SSN feeding support is crucial. The implementation of these strategies for sustainable change requires collaboration among facility staff, administrators, regional and local health department leaders, and non-governmental stakeholders involved in maternal and neonatal health.

## ACKNOWLEDGEMENTS

The authors would like to express their gratitude to the participating clinicians and healthcare facilities, the Amhara Regional Health Bureau, the Amhara Public Health Institute, and the Emory Ethiopia Bahir Dar staff for their invaluable support and collaboration.

## ETHICAL STATEMENT

The Research Ethics Committee (REC) of the Amhara Public Health Institute (APHI) approved the study (Reference Number: NoH/R/T/T/D/5/9, Date:12/13/2021). The Institutional Review Board of Emory University granted an exemption for this research study as it falls under the category of non-human subject research.

## AUTHOR CONTRIBUTIONS

Conceptualization; AG, JC, MY

Data curation; YAT, DA, MA

Formal analysis; YAT, MCE

Funding acquisition; AG, JC, MY

Investigation; YAT, AG, DA, JC, MA, MY

Methodology; AG, JC, MY, YAT

Project administration; MLB, HB, YAT

Resources; Software; Supervision; Validation; Visualization AG, JC, MY, YAT

Roles/Writing – original draft; and Writing – review & editing YAT, ZT, AG, MY, JC

## DATA AVAILABILITY

The data are available upon reasonable request to the authors.

